# Unsupervised Learning for Large Scale Data: The ATHLOS Project

**DOI:** 10.1101/2021.04.01.21254751

**Authors:** Petros Barmpas, Sotiris Tasoulis, Aristidis G. Vrahatis, Panagiotis Anagnostou, Spiros Georgakopoulos, Matthew Prina, José Luis Ayuso-Mateos, Jerome Bickenbach, Ivet Bayes, Martin Bobak, Francisco Félix Caballero, Somnath Chatterji, Laia Egea-Cortés, Esther García-Esquinas, Matilde Leonardi, Seppo Koskinen, Ilona Koupil, Andrzej Pająk, Martin Prince, Warren Sanderson, Sergei Scherbov, Abdonas Tamosiunas, Aleksander Galas, Josep MariaHaro, Albert Sanchez-Niubo, Vassilis P. Plagianakos, Demosthenes Panagiotakos

## Abstract

Recent technological advancements in various domains, such as the biomedical and health, offer a plethora of big data for analysis. Part of this data pool is the experimental studies that record various and several features for each instance. It creates datasets having very high dimensionality with mixed data types, with both numerical and categorical variables. On the other hand, unsupervised learning has shown to be able to assist in high-dimensional data, allowing the discovery of unknown patterns through clustering, visualization, dimensionality reduction, and in some cases, their combination. This work highlights unsupervised learning methodologies for large-scale, high-dimensional data, providing the potential of a unified framework that combines the knowledge retrieved from clustering and visualization. The main purpose is to uncover hidden patterns in a high-dimensional mixed dataset, which we achieve through our application in a complex, real-world dataset. The experimental analysis indicates the existence of notable information exposing the usefulness of the utilized methodological framework for similar high-dimensional and mixed, real-world applications.

## 2 Introduction

In recent years, the embracement of technology in various domains has experienced remarkable growth, and as a result, the data generation has increased significantly and is expected to multiply in the near future (Alharthi, Krotov and Bowman 2017) (Gupta and Rani 2019) (Gu, et al. 2017). Simultaneously, utilizing the innovations in the collection, recording, and storage, huge databases are being constructed, including various data types from different sources (Roh, Heo and Whang 2019) (Plageras, et al. 2018) (Wang, Ng and Brook 2020). However, along with the vast increase in size, it is not easy to simultaneously produce and categorize them, thus leading to heterogeneity phenomena. Therefore, advanced processing techniques that allow the management of such data become necessary, aiming to retrieve patterns, reduce dimensionality and at the same time automate the processing (Hsu and Glass 2018) (Usama, et al. 2019) (Wang, et al. 2019). This development constantly reinforces the “Big Data” term, enabling the research community to enhance insight, decision-making, and process automation while simultaneously necessitating cost-effective, novel means of information processing (Sagiroglu and Sinanc 2013). The arising Big Data characteristics though may weaken prediction accuracy and pattern discovery by imposing noise through measurement errors and false correlations (Fan, Han and Liu 2014).

Diverse dimensionalities and heterogeneous structure are two key features that intensify large-scaled data complexity. When we consider massive heterogeneous data (Zhu, et al. 2018), the features may represent different types of information of the same individual (Fan and Fan 2008). These issues become significant challenges when trying to enable data aggregation (Fan and Fan 2008) (Fan, Guo and Hao 2012) (Li, et al. 2016).

In an early stage of data centralized information systems, the focus is on finding the best feature values to represent each observation (Tufekci 2014) (Keyes and Westreich 2019). This type of sample feature representation inherently treats each individual as an independent entity without considering their social connections. In a dynamic world, the features used to represent the individuals and the social ties used to represent their connections may also evolve concerning temporal, spatial, and other factors. Such complexity is becoming part of Big Data applications’ reality, introducing significant computational challenges for data analytics (Jin, et al. 2015) (Marx 2013).

Nevertheless, techniques and methods that fall into the category of unsupervised learning have shown encouraging results and are able to overcome some of the difficulties of vast, heterogeneous data (Casolla, et al. 2019) (Hameed, et al. 2018) (Ma, et al. 2017) (Xiang, et al. 2018). In this study, we focus on the area of unsupervised learning, presenting a complete methodological procedure that utilizes recent advances in the field. We begin with a review of state of the art methods for clustering and dimensionality reduction and conclude with their utilization in a real world dataset, characterized by the aforementioned challenges. The organization of this paper is as follows. Chapter 3 presents the basic principles and characteristics of unsupervised learning and introduces some indicative techniques for two subcategories, dimensional reduction, and clustering respectivelly. Chapter 4 presents the methodology followed for the experimental procedure, in an attempt to extract knowledge from the data set at hand, that of the ATHLOS Cohort. Finally, in chapter 5, we conclude with a discussion upon the results and potential perspectives.

## 3 Unsupervised Learning Methods as an Approach for Knowledge Extraction

Unsupervised learning is one of the main categories of Machine Learning, along with supervised and reinforcement learning (Hinton, Sejnowski, et al. 1999), and hybrid methods like semi-supervised learning (Zhu and Goldberg 2009) (X. J. Zhu 2005). Unsupervised learning can be divided into three main application fields (Ghahramani 2003). The first one concerns data samples segmentation by some shared attributes, next is the outlier detection (Both can be attributed to Clustering methods (Jiang and An 2008) (Chawla and Gionis 2013)), while the last is dataset simplification by aggregating variables with similar attributes, a procedure known as Dimensionality Reduction accompanied with Feature Selection (Wei and Billings 2006) (Masaeli, Fung and Dy 2010) (Mladenič 2005). In summary, Unsupervised Learning aims to study the intrinsic structure of the data to find patterns that should not be considered plain, unstructured noise (Ghahramani 2003).

Each of the subcategories has the potential to extract helpful information regarding a dataset. However, the combination of them has been previously shown to produce encouraging results (Diaz-Papkovich, Anderson-Trocmé and Gravel 2019) (Allaoui, Kherfi and Cheriet 2020) (Hozumi, et al. 2021). In what follows, we highlight the some of most representative techniques from each subcategory that we also utilize on our experimental analysis. Our aim is to incorporate both well-established and recent state-of-the-art and popular methods.

### 3.1 Dimensionality Reduction for Pattern Discovery through Visualization

Biomedical and health technologies are constantly evolving generating ultra-high dimensional data since we have several features for each record. Sampling techniques aim to reduce the dataset’s size but still do not offer a solution for high-dimensional datasets. In such cases, Dimensionality Reduction precedes Clustering procedures as a preprocessing step (Kaski 1998) (Yan, et al. 2006). Dimensionality Reduction (DR) aims to solve the Curse of Dimensionality (Bellman n.d.) depicting that when the dimensionality increases, the volume of the space increases at such a rate that the dataset becomes sparse, opposing statistical methods. The goal is to find low-dimensional representations of the data that retain their fundamental properties, typically in two or three dimensions (Ghodsi 2006) (Sorzano, Vargas and Montano 2014). As such this process is also essential for data visualization in lower dimensions (Xia, et al. 2017). Visualization tools can assist in identifying the data structure while plotting the data in two dimensions allows researchers to pinpoint any remaining technical variability source between samples, which should be removed by normalization (Rostom, et al. 2017).

Meanwhile, well-established visualization techniques that have been proved effective for small or intermediate size data face a significant challenge when applied to big and high dimensional data. Visualizing high-dimensional data could allow the discovery of hidden relationships between the hidden variables and numeric values (Xia, et al. 2017). Although there is remarkable progress in this field, identifying an extremely low-dimensional representation of large-scale and high-dimensional data remains a major challenge. Dimensionality reduction techniques able to handle large data are presented below, either traditional, established methods or state-of-the-art approaches specifically designed for Big Data scenarios.

Principal component analysis (PCA) is probably one of the most popular multivariate statistical technique used by almost all disciplines. It is also likely the oldest multi-variable technique. Its origins date back to Pearson (Pearson 1901). PCA is a statistical process that uses a rectangular transformation to convert a set of observations of possible associated variables (entities each receiving different numeric values) into a set of values of linearly unrelated variables called main variables. This transformation is defined so that the first principal component has the highest possible variance (i.e., gives the highest volatility to the data), and each subsequent one has the next highest variance, subject to the constraint that it is rectangular with the previous components. By visualizing the two main components, the user can apprehend some of the topologies that the data have while also being assured that most of the relevant information is still preserved. PCA, though, is mainly able to produce acceptable results in linear datasets (Shah, et al. 2013). For this reason, a large number of non-linear dimensionality reduction techniques have been created to preserve the topology of the dataset better. In what follows, we present some state-of-the-art tools that have seen adoption in recent years.

Van der Maaten and Hinton proposed t-distributed stochastic neighborhood embedding (t-SNE) in 2008. Until recently, it was considered to have vast applicability and great accuracy (Kobak and Berens 2019) (Rauber, et al. 2016). This technique is an extension of the SNE as proposed by Hinton and Roweis in 2003 (Hinton and Roweis, Stochastic neighbor embedding 2003), which is a technique that minimizes the Kullback-Leibler (Kullback 1997) deviation of the scaled similarities among pairs of points both in high and low dimensional spaces. SNE uses a Gaussian kernel to compute similarities in a high and low dimensional space. The t-Distributed Stochastic Neighborhood Embedding improves SNE by using a t-Distribution as a kernel in low dimensional space. Because of the heavy-tailed t-distribution, t-SNE maintains local neighborhoods of the data better and penalizes wrong embeddings of dissimilar points (Maaten and Hinton 2008). This property makes it especially suitable to represent clustered data and complex structures in a few dimensions. The minimization of the Kullback-Leibler divergence with respect to the points is performed using gradient descent.

Uniform Manifold Approximation and Projection (UMAP) (McInnes, Healy and Melville 2018) is a new multi-dimensional learning technique for non-linear manifolds. The UMAP algorithm is competitive with t-SNE in terms of visualization quality and, according to the authors, maintains a more comprehensive structure with superior runtime performance. Furthermore, UMAP has no computational restrictions on the embedding dimension, making it viable as a general-purpose dimension reduction technique for machine learning. What characterizes UMAP is that it uses local approximations of the dataset and then links them with fuzzy unions to construct simplicial sets representing the high-dimensional data’s topological geometry.

LargeVis (Tang, et al. 2016) is another novel visualization technique. Many recent approaches like the t-SNE, as mentioned above, construct a K-nearest neighbor graph and then project the graph into the 2-d space. LargeVis follows a similar procedure. First, LargeVis produces an accurately approximated K-nearest neighbor graph from the data and then layouts the graph in the low-dimensional space but, in contrast, uses an efficient algorithm for K-nearest neighbor graph construction and a principled probabilistic model for graph visualization. The whole procedure thus could scale to millions of high-dimensional data points. According to the authors, LargeVis outperforms state-of-the-art methods in both efficiency and effectiveness.

### 3.2 Unsupervised Learning through Clustering

Broadly defined clustering aims to identify subgroups (clusters) in the data that are distinguished by an appropriate measure of similarity (or regularity), without any previous knowledge about the assignment of observations to clusters or even the presence of clusters (Everitt, et al. 2011). The main goal is to group sets of objects so that samples in the same group are more similar to each other than samples in different groups. Clustering is among the most used exploratory data analysis techniques (Berkhin 2006) (Gan, Ma and Wu 2020) (Jajuga, Sokolowski and Bock 2012). The recent explosion of data availability leads to an ever-growing tendency to “let the data speak” (Cios, Pedrycz and Swiniarski 1998). However, the properties of novel data sources and the increasing size, dimensionality, and speed at which data are captured pose challenges for established methods. Applications for cluster analysis include gene sequence analysis, market research, and object recognition. In general, clustering techniques aimed at big data can be categorized into single-machine and distributed clustering algorithms (Shirkhorshidi, et al. 2014).

Partitioning clustering algorithms aim to divide the space that the data points lie on into sub-spaces that each contains a set of data points, according to a pre-specified number. Their simplicity and performance have attracted the research community’s interest even in recent years proposing sophisticated variations and big data capable versions (Sreedhar, Kasiviswanath and Reddy 2017). One significant advantage of such approaches is that every set of data points has a distinct center (representative) As a result, a new point can be efficiently assigned to the appropriate set after the fact. Usually, Partitioning methods (K-means, PAM clustering) are most suitable for finding spherical or convex clusters, meaning they work well only for compact and well-separated clusters. Moreover, they can be severely affected by the presence of noise and outliers in the data. For such cases, density-based approaches are usually employed (Gao, et al. 2016) (Hahsler, et al. 2017); however, they typically fall short in the presence of vast amounts of high dimensional data. The computational and memory requirements in Big Data scenarios are prohibitive for density-based algorithms. Simultaneously, we lose the ability to extract representatives that allow a straightforward procedure of allocating new samples into clusters.

Kmeans (Forgy 1965) algorithm is an iterative algorithm that tries to partition a dataset in *K* pre-defined discrete non-overlapping clusters where each data point belongs to only one group. K-means is a simple algorithm that has been used in a variety of fields. The goal is to make the data points belonging to the same cluster as similar as possible while keeping the clusters as distant from each other as possible. For a set of samples (*x*1, *x*2, …, *xn*) in the d-dimensional space, cluster the dataset into clusters *C* = {*C*1, *C*2, …, *Ck*} by minimizing the within-cluster sum of squares or variance. The algorithm assigns data points to a cluster in such a way that the total sum of the squared distances among the data points and their cluster’s centroid (mean value of the data points that belong to a cluster) is minimized. The less variation there is within groups, the more similar data points are within each cluster.

Hierarchical clustering constructs hierarchies of clusters in a top-down (agglomerative) or bottom-up (divisive) fashion. The former starts from n clusters, where n stands for the number of data points, each containing a single data point and iteratively merging the clusters satisfying certain closeness measures. Divisive algorithms follow a reverse approach, starting with a single cluster containing all the data points and iteratively split existing clusters into subsets. Hierarchical clustering algorithms have been shown to result in high-quality partitions, especially for applications involving clustering text collections. Nonetheless, their high computational requirements usually prevent their usage in big data scenarios. However, more recent advancements in both agglomerative (Murtagh and Legendre 2014) (Zhang, Zhao and Wang 2013) and divisive strategies (Sharma, López and Tsunoda 2017) (Tasoulis, et al. 2014) have exposed their broad applicability and robustness. In particular, when divisive clustering is combined with dimensionality reduction (Hofmeyr 2016) (Pavlidis, Hofmeyr and Tasoulis 2016), we can still get methods capable of indexing large data collection that allow fast sample allocation due to their tree structure.

The Normalized Cut Divisive Clustering (Ncutdc) (Hofmeyr 2016) algorithm is a computationally efficient divisive clustering algorithm relying on hyperplane separators. It generates a binary partitioning tree by recursively partitioning a dataset using a hierarchical collection of hyperplanes with low normalized cut measured across them. As in the minimum density hyperplane case, the projection pursuit problem is formulated as a minimization problem. The normalized cut (NCut) (Shi and Malik 2000) associated with a partition of *X* into clusters *C*1, …, *Ck* is expressed as,

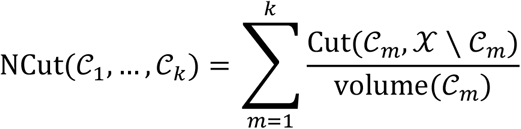

By minimizing the normalized cut, this leads to solutions for which *Cut(cm,X \ Cm)* is small and volume (*Cm*) is large, for all *m*. Since 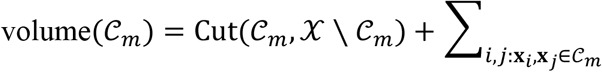 similarity(*x*_*i*_,*x*_*j*_), where the last term is the total internal similarity of points in *C*_*m*_ this implies that the similarity within clusters is high whereas the similarity between clusters is low. However, the NCut problem is NP-hard, and, instead, a continuous relaxation of the problem known as spectral clustering (Shi and Malik 2000) (Von Luxburg 2007) is considered. This leads to a reduction in complexity, but this method remains applicable only to moderate size situations.

The Genie (Gagolewski, Bartoszuk and Cena 2016) algorithm is an alternative for the more classical, single-linkage criteria hierarchical clustering. The algorithm aims to offset the disadvantages of the single linkage scheme, that is, sensitivity to outliers, the creation of very skewed dendrograms, and consequently not reflecting the actual underlying data structure unless there are well-separated clusters. Simultaneously, to retain the single-linkage’s simplicity and efficiency, the following linkage criterion, referred to as the Genie algorithm, was created. Let F be a fixed inequity measure (e.g., the Gini-index) and *g* ∈ (0, 1] be some threshold. At step j: If 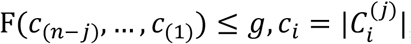, apply the original single linkage criterion, otherwise, if F(*c*_(*n*-*j*)_, …, *c*_(1)_) > *g*, restrict the search domain only to pairs of clusters such that one of them is the smallest. This modification prevents extreme increases of the chosen inequity measure and forces early merges of small clusters with others.

Finally, in density-based clustering, a cluster is a set of data objects spread in the data space over a contiguous region of high density of objects. Density-based clusters are separated from each other by contiguous regions of low density of objects. Data objects located in low-density regions are typically considered noise or outliers. Density-based clustering algorithms are able to discover arbitrary-shaped clusters, but usually suffer from increased computational costs, preventing them to be scalable. DensityPeaks (Rodriguez and Laio 2014) algorithm is a novel density-based approach. Similar to the K-medoids method, it has its basis only in the distance between data points. Like DBSCAN (Hahsler, et al. 2017) and the mean-shift process, it can detect non-spherical clusters and automatically find the correct number of clusters. As in the mean-shift method, the cluster centers are defined as local maxima in the density of data points. However, unlike the mean-shift method, this procedure does not require embedding the data in a vector space and maximizing explicitly the density field for each data point. The algorithm assumes that cluster centers are surrounded by regions with lower local density and are relatively far away from higher local density points. For each data point, the algorithm computes two measures: its local density and its distance from samples of higher density. Both these measures depend exclusively on the intervals between data points, which are considered to satisfy the triangular inequality.

## 4 Experimental Analysis

To the best of the writer’s knowledge, there is no singular unsupervised methodology that is able to extract all the available information from any dataset (Adam, et al. 2019). In many cases, methods from different fields are combined, resulting in more sophisticated techniques with benefits from all the components. This section presents some combinatorial methodologies and an example of such schemes in a Big Data scenario. More precisely, we will implement a complete unsupervised learning methodology for the ATHLOS cohort dataset while also utilizing very recent techniques for variable exploration.

### 4.1 Data Specification and Pre-processing

ATHLOS (Ageing Trajectories of Health: Longitudinal Opportunities and Synergies) is a project funded by the European Union’s Horizon 2020 Research and Innovation Program, which aims to interpret aging’s impact on health better. The ATHLOS project provides a harmonized dataset (Sanchez-Niubo, et al. 2019), built upon several longitudinal studies and originated from five continents. More specifically, it contains samples coming from more than 355,000 individuals who participated in 17 general population longitudinal studies in 38 countries. Based on the WHO healthy aging framework, researchers from the ATHLOS consortium reviewed measures of functional ability in the aging cohorts and identified 47 items related to health, physical, and cognitive functioning. The consortium harmonized these 47 items into binary variables and used item-response theory modeling to generate a common measure for healthy aging across cohorts.

In this paper, we used 15 of these studies, namely the 10/66 Dementia Research Group Population-Based Cohort Study (Prina, et al. 2017), the Australian Longitudinal Study of Aging (ALSA) (Luszcz, et al. 2016), the Collaborative Research on Ageing in Europe (COURAGE) (Leonardi, et al. 2014),the ELSA (Steptoe, et al. 2013), the study on Cardiovascular Health, Nutrition and Frailty in Older Adults in Spain (ENRICA) (Rodríguez-Artalejo, et al. 2011), the Health, Alcohol and Psychosocial factors in Eastern Europe Study (HAPIEE) (Peasey, et al. 2006), the Health 2000/2011 Survey (Koskinen 2018), the HRS (Sonnega, et al. 2014), the JSTAR (Ichimura, Shimizutani and Hashimoto 2009), the KLOSA (Park, et al. 2007), the MHAS (Wong, Michaels-Obregon and Palloni 2017), the SAGE (Kowal, et al. 2012), SHARE (Börsch-Supan, et al. 2013), the Irish Longitudinal Study of Ageing (TILDA) (Whelan and Savva 2013) and the Longitudinal Aging Study in India (LASI) (Arokiasamy, et al. 2012). The 15 general population longitudinal studies utilized in this work consist of 990,000 samples in total, characterized by 184 variables. The version used here is a preprocessed dataset where a selection of variables has been removed along with several samples as described in (Anagnostou, et al. 2021). The resulting data matrix constituted by 770,764 samples and 107 variables has been imputed using the Vtreat (Zumel and Mount 2016) imputation method in order to populate the missing values in a meaningful manner. As a result 458 dummy variables has been created constituting the final data dimensionality.

### 4.2 Data Visualization for Pattern Recognition

To this end, we employ a series of dimensionality reduction algorithms for embedding the ATHLOS dataset in two dimensions. All methodologies are implemented using the R-project open-source environment for statistical computing, and experiments were conducted for each algorithm to tune its hyper-parameters. In what follows, we provide the details of each implementation tested, referring to the corresponding R packages when possible. For PCA, we employed the implementation found in the “dimRed” package (Kraemer, Reichstein and Mahecha 2018). The fast “Rtsne” implementation for tSNE (Krijthe 2015) was used, and the two main hyper-parameters were set as follows, “perplexity” at a range of 30 to 800 and “theta” at a range of 0.1 to 1. The “uwot” implementation of the UMAP algorithm (McInnes, Healy and Melville 2018) is used, and the two main hyper-parameters were examined, “cluster neighbors” at a range from 15 to 100 and “minimum distance” at a range from 0.01 to 0.15. Finally, the “largeVis” implementation of the LargeVis algorithm (Elberg 2020) was used, and the hyper-parameters, “Kapa” and “max iterations,” were set at a range from 10 to 200 and from 10 to 50 accordingly.

In Figure 4.1, we observe the resulting visualization for all methods across the samples. It is evident that PCA tends to produce a more coherent representation without however distinguishing any particular clusters. The advantage of this technique, however, is that the coordinates have a strict and valuable definition. UMAP and LargeVis seem to produce very similar results creating several very distinct clusters. tSNE also created groups that were, however, larger, taking into account their in-between cluster distances. As depicted in Figure 4.1, the non-linear dimensionality reduction techniques, in this case, tend to separate more clearly groups of individuals that lie more closely in the intrinsic dimensionality manifold.

**Figure 4.1:**
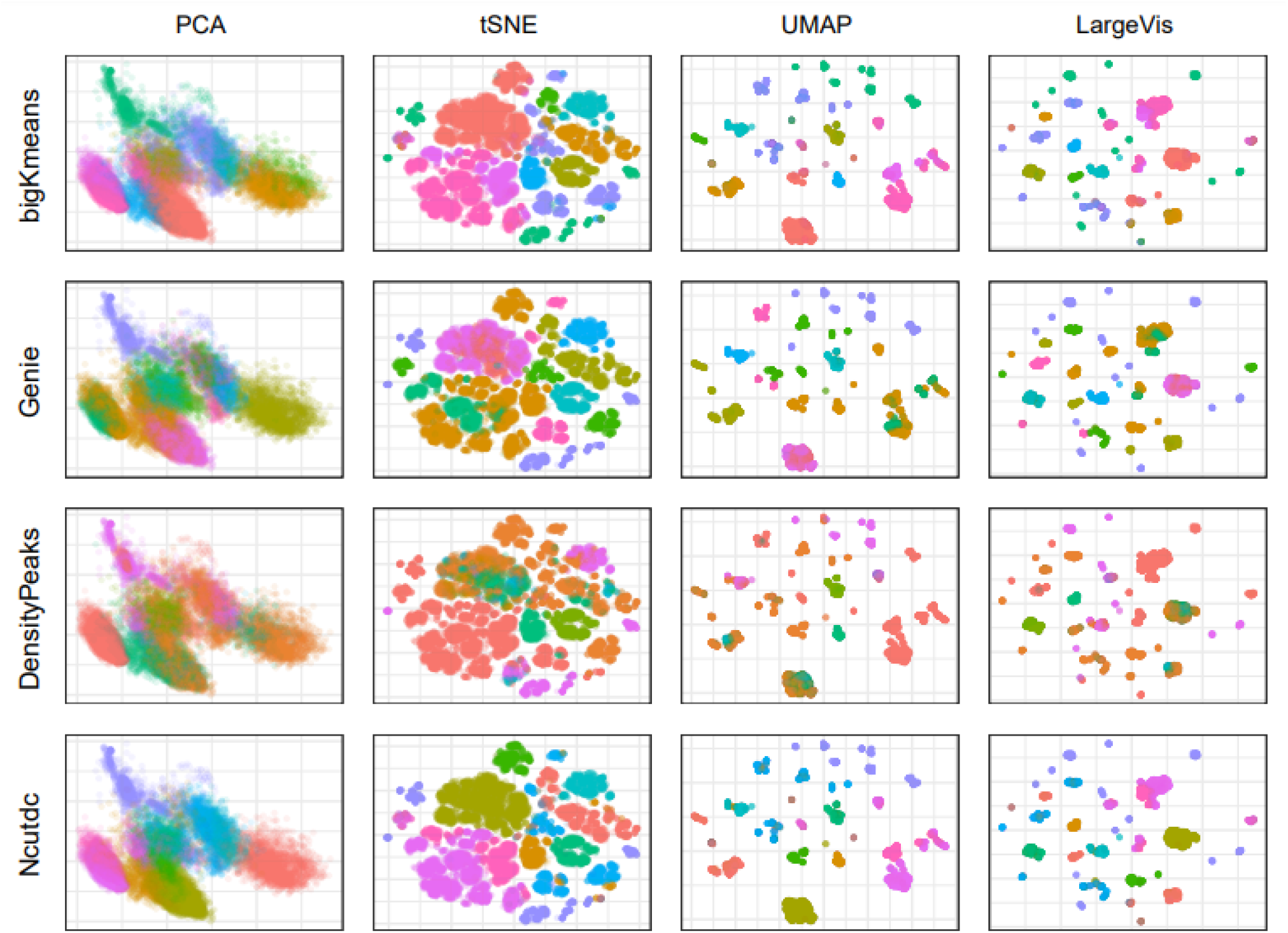
2D visualization of PCA, tSNE, UMAP and LargeVis embeddings (column-wise) on the ATHLOS dataset clustered with “bigKmeans”, “Genie”, “DensityPeaks”, and “Ncutdc” clustering algorithms (row-wise) accordingly. Circular points represent each sample of data colored by the corresponding cluster ID resulting from the respective clustering algorithms.

### 4.3 Clustering for Verification

The next step of our analysis is to determine the existence of clusters that could be verified through visualization. As a first step, we looked at the clustering tendency of the dataset. That is whether the data contain any inherent grouping structure. For this purpose, we calculated the well-known “Hopkins” statistic (Lawson and Jurs 1990), for which values close to 1 indicate a clusterable dataset. In our case, using the “clusterland” R package (YiLan and RuTong 2015)(, the corresponding calculated value is *83%*, suggesting a high degree of clusterability. Subsequently, for brevity and the sake of the reader’s convenience, we choose to determine a representative number of clusters to accompany visualizations instead of providing extensive parameter analysis, which would hinder visual interpretation. Apparently, this task is not trivial, and there are over thirty different approaches in the literature regarding this exact case. After both heuristic experiments and counseling the results provided by the “factoextra” R package (Kassambara, Mundt and others 2017), we propose that any number between *4* to *12* clusters is an appropriate choice. Consequently, we proceed to the clustering step. For the Kmeans algorithm, we chose the memory efficient implementation found in (Emerson and Kane 2020), optimized for large scale applications. For the Genie algorithm, the corresponding R package “genie” (Gagolewski, Bartoszuk and Cena 2016) was used. For the Ncutdc algorithm, the implementation found in the authors ‘s “PPCI” (Hofmeyr and Pavlidis, PPCI: an R Package for Cluster Identification using Projection Pursuit 2019) package was used. Lastly, any attempt to use popular density-based approaches in the original dimension space, unfortunately, failed due to the dataset’s scale. In particular, we employed the recent implementation of the DensityPeaks algorithm found in (Pedersen, Hughes and Qiu 2017) but hardware requirements exceeded 1TB of RAM usage. Thus, we chose to firstly reduce the number of dimensions down to 50 with the PCA method in order to be able to run all the algorithms in a reasonable amount of time. Afterward, we extracted a uniformly random subsample of the dataset of 20000 samples. The DensityPeaks algorithm lacks the ability to tune the parameters “rho” and “delta” that affect the number of retrieved clusters. Instead, provided is a graphical tool with which the user can manua**l** y set the respective values through a scatter plot’s visual investigation. In our tests, we set these as the mean values across the parameters calculated for all samples.

Cluster memberships depicted in Figure 4.1 arise from the application of bigKmeans, Genie, DensityPeaks and Ncutdc clustering algorithms on the ATHLOS dataset in the 50-d dimensional space, with k=10. It is observed that there is a clear correlation between the embeddings in the two-dimensional space and the clusters assigned by bigKmeans. Also, the Ncutdc algorithm produces very distinct clusters, similar to bigKmeans, with minor tangling, as it is the case of Genie. Lastly, for DensityPeaks, a large number of data points appear to be mixed regarding the allocated cluster and the corresponding location within the embedding. Thus, this result further verifies that the visualizations were representative and that there are groups of individuals with some distinct characteristics. However, which variables are the dominant separation attributes is not easy to interpret only with statistical measures or exhaustive search for each feature in every cluster.

To numerically validate the clustering result of the aforementioned algorithms, we employ an implementation of the Silhouette index (Rousseeuw 1987) Dunn index (Bezdek and Pal 1995) and Separation index specifically designed for Large scale data provided by the “fpc” R package (Hennig 2020). Silhouette is a method of interpretation and validation of consistency within a cluster. The silhouette index compares how similar an object is to its group compared to other clusters. The silhouette has a range of -*1* to *+1*, where a high value indicates that the object is well matched to its cluster and poorly matched to neighboring clusters. The Dunn index depicts the ratio of the smallest distance between observations not in the same cluster to the largest intra-cluster distance and has a value between zero and infinity. The Separation Index is defined based on the distances for every data point to the closest neighbor not in the same group. The separation index then depicts the mean of the smallest proportion of these distances. This allows formalizing separation less sensitive to a single or a few ambiguous points. Considering the results of the aforementioned metrics presented in Table 1, it is verified that “DensityPeaks” do not separate clusters as effectively as the other three methods with “Ncustdc” achieving the highest scores.

**Table 1:** Internal Validation metrics for the Clustering algorithms. Larger values depict grater similarities within the same cluster, while the clusters been more separate.

|  | **Silhouette** | **Dunn** | **Separation** |
| --- | --- | --- | --- |
| *BigKmeans* | 0.1265699 | 0.738058 | 8.343318 |
| *Genie* | 0.04942602 | 0.7035942 | 6.956037 |
| *DensityPeaks* | -0.1252029 | 0.4331565 | 8.033905 |
| *Ncutdc* | **0.1301279** | **0.7423549** | **8.436816** |

In an attempt to extract knowledge based on the identified patterns, the 2d embedding of the tSNE method was colored according to some categorical variables of interest (see Figure 4.2). The variables were selected according to the following scheme. For every categorical variable, we calculated the Silhouette (Rousseeuw 1987), Dunn (Bezdek and Pal 1995) and Separation (Hennig 2020) indexes for the 2d embeddings, considering the variables as a typical clustering results. Then, we chose six variables that presented the highest separability according to these metrics. There are apparent correlations with some of the variables and the resulting clumps in the visualization, implying that individuals have some distinct characteristics. We interestingly observe that we are able to identify separable regions characterized by particular levels of the categorical variables, potentially leading to straight forward cluster characterization.

**Figure 4.2:**
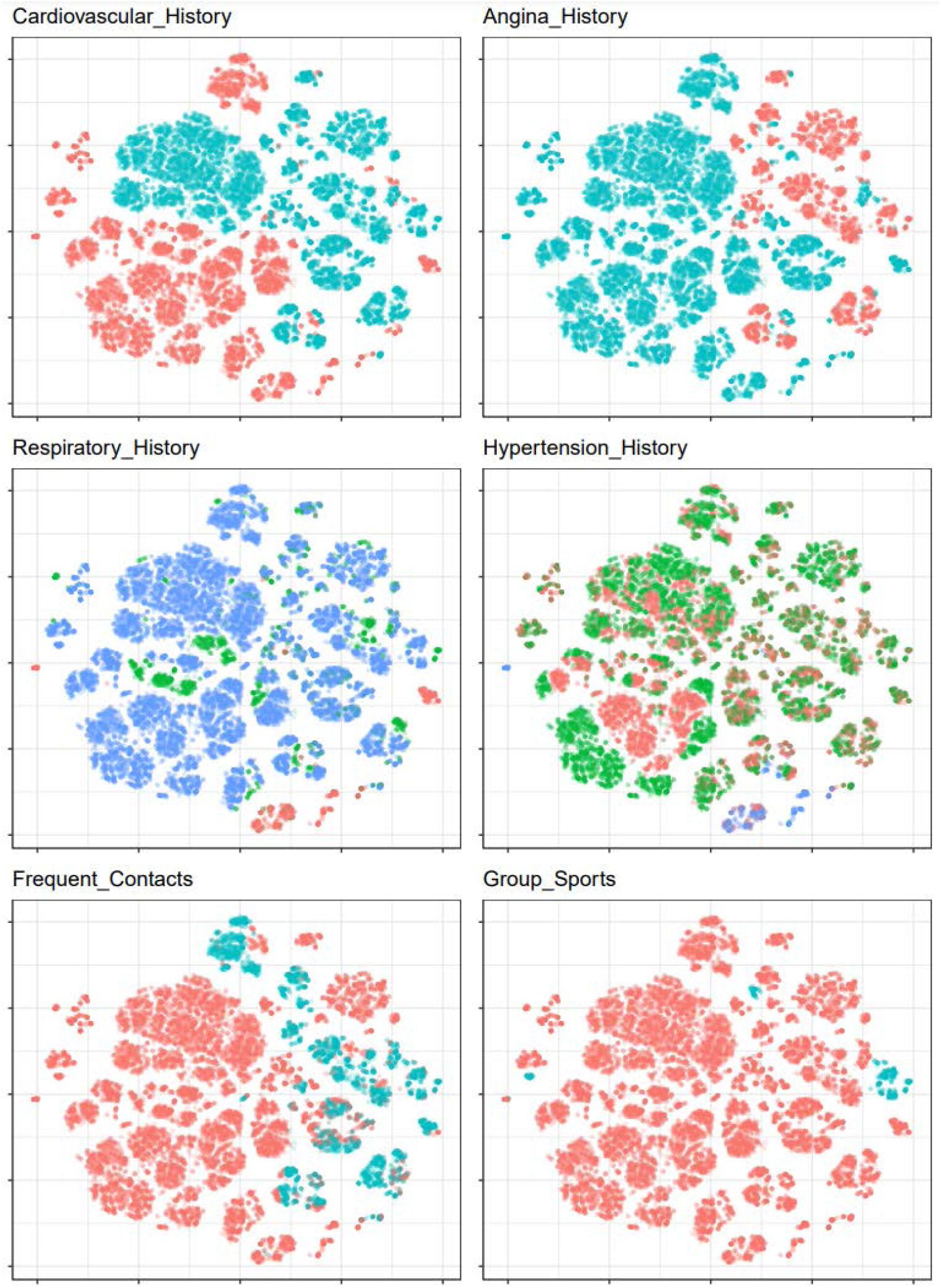
Visualizations of the 2d embedded dataset using t-SNE with respect to a selection of variables found in the ATHLOS dataset. Each subplot corresponds to different variables, while different colors correspond to different values for every variable.

In more detail, the features depicted in Table 2 are the following: Cardiovascular_History: Is a binary variable and refers to the history of stroke or myocardial infarction (heart attack) of an individual. Angina_History: Is a binary variable and refers to the “h_angina” variable of the original dataset that depicts whether or not an individual had a history of angina. Respiratory_History: Refers to the categorical “h_respiratory” variable of the original dataset that depicts one’s history of chronic respiratory diseases such asthma, CPD, COPD, bronchitis, etc. This variable is then transformed with the “ Vtreat” package to an encoding that expresses the within-group deviation of the outcome conditioned on each categorical level in the original data and thus, amend the high cardinality of the variable. Hypertension_History: Refers to the categorical “h_hypertension” variable of the original dataset that depicts one’s history of hypertension for an individual. This variable is then transformed with the “Vtreat” package to an encoding that expresses the within-group deviation of the outcome conditioned on each categorical level in the original data and thus, amend the high cardinality of the variable. Frequent_Contacts: Is a binary variable and refers to the “cont_fr” variable of the original dataset that depicts if an individual has frequent contacts with friends/neighbours. Group_Sports: Is a binary variable and refers to the “sport” variable of the original dataset that depicts if an individual participates currently in group sport activities.

**Table 2:** Internal Validation metrics for the most separated features. Larger values depict grater similarities within the samples of with the same value for that particular feature, while the samples not in the same category been more separate.

|  | **Silhouette** | **Dunn** | **Separation** |
| --- | --- | --- | --- |
| *Cardiovascular_History* | 0.2239903 | 1.2507654 | 1.1195133 |
| *Angina_History* | 0.2198167 | 1.2698951 | 1.2975713 |
| *Respiratory_History* | 0.2059388 | 1.3212471 | 1.0268838 |
| *Hypertension_History* | 0.1861763 | 1.2968042 | 0.9150790 |
| *Frequent_Contacts* | 0.1800876 | 1.2259719 | 0.1931220 |
| *Group_Sports* | 0.1533813 | 1.2334624 | 4.9907552 |

### 4.4 Variable Importance through Heatmaps

Motivated by the previous sections’ findings, we introduce an additional step to visualize the regions’ variable differences. For the final step, we chose to include a novel method so far utilized only for Genomics in Gene Expression (Linderman, Rachh, et al. 2019). The process incorporates both tSNE and clustering in order to produce a Heatmap that visualizes many variables (instead of genes) of interest at the same time. To build the t-SNE Heatmap introduced in (Linderman, Rachh, et al. 2019), we initially compute t-SNE embeddings of the Variables of interest into one dimension. This implementation incorporates the FIt-SNE (Linderman, Rachh, et al. 2017), which is scalable to millions of Variables in terms of computational time. Then, the 1D t-SNE embeddings are discretized in 100 bins, and the representation of each variable is produced by the sum of its expression in the samples contained in each bin, while each variable corresponds to a vector in R^100. Hierarchical clustering upon the aforementioned vectors produces even more meaningful results. Subsequently, for some set of variables of interest, the algorithm “enriches” the variables that have a similar expression pattern in the t-SNE (see Figure 4.3). Afterward, these vectors are transformed into Heatmap format, with each row being a variable and each column a bin using the heatmaply R package (Galili, et al. 2018).

**Figure 4.3:**
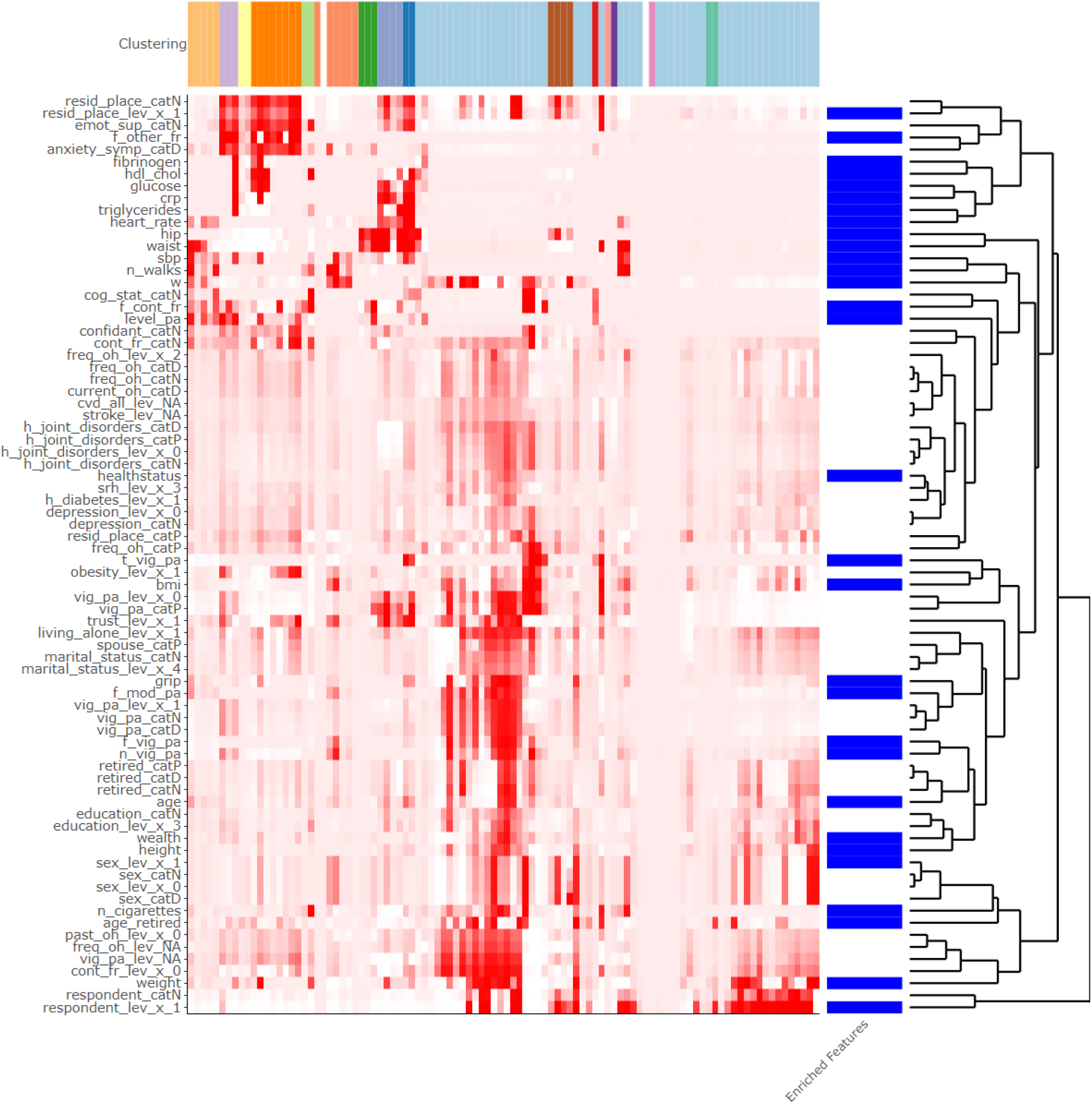
Heatmap Visualization of the Athlos dataset. Depicted are the Variables in every row, colored according to their respective value, with red depicting a larger value. The columns represent 100 bins of samples and are arranged according to their 1d-tSNE embedding. Also, the “Clustering” bar depict the cluster assignment of the DBSCAN algorithm run in the 2d-tsne embedding of the dataset. The “Enriched_Features” bar depicts the additional variables returned by the 1d-tsne-heatmap function where in blue color are the variables of interest as an input for the function and with white color are the additional variables returned for the “enrich” parameter equal to 5.

This is possible because it has been previously shown that t-SNE preserves the cluster structure of well-clustered data regardless of the embedding dimension (Linderman, Rachh, et al. 2019), and thus 1D t-SNEs contains the same information as 2D t-SNEs. The resulting vectors visualized in the produced Heatmap presented in Figure 4.3 provides a clear depiction of the variables’ behavior among the clusters, with hundreds of variables visualized at the same time.

## 5 Conclusions

This study provided insight into recent Unsupervised Learning methods and their popular implementations through their application on a real word complex dataset. A shown, based on these methods we were able to provide a comprehensive example of knowledge extraction and pattern recognition analysis. In addition, we utilized a promising novel unsupervised learning approach from the Gene Expression field to provide useful information of Variable expression for the ATHLOS cohort dataset at hand, exposing the method’s usefulness in similar Big Data tasks.

## Data Availability

Data sharing is not applicable to this article

## 6 Acknowledgements

This work is supported by the ATHLOS (Aging Trajectories of Health: Longitudinal Opportunities and Synergies) project, funded by the European Union’s Horizon 2020 Research and Innovation Program under grant agreement number 635316.

